# Feasibility and Validity of Clinical Outcome Measures in a Prospective Natural History Study of *STXBP1*-Related Disorders

**DOI:** 10.64898/2026.09.15.26361330

**Authors:** Samuel R. Pierce, Julie M. Orlando, Sarah M. Ruggiero, Ian McSalley, Kristin Cunningham, JoeyLynn Coyne, Sarah Tefft, Jillian L. McKee, Ingo Helbig, Bintou Bane, Torrey Chisari, Anna Prentice, Rency Dhaduk, Carlyn Glatts, Charlene Woo, Maya G. Mosner, Stephanie A. Zbikowski, Zachary Grinspan, Juliet Knowles, Hsiao-Tuan Chao, Katherine Xiong, Elizabeth Berry-Kravis, Sepideh Tabarestani, Charlene Son Rigby, James Goss, Megan Abbott, Scott Demarest, Andrea Miele, Benjamin Prosser, Michael J. Boland

## Abstract

**Aims:** To describe the developmental phenotype of individuals with *STXBP1*-related disorders (*STXBP1*-RD) in a prospective natural history study using standardized outcome measures.

**Methods:** Data were collected from 155 individuals with a mean age of 8.2 years. The Bayley Scales of Infant and Toddler Development-Fourth Edition (Bayley-4), Hand Manipulation and Eye Hand Coordination subtests of the Peabody Developmental Motor Scales-Third Edition (PDMS-3), and Gross Motor Function Measure-66 (GMFM-66) were assessed. The Gross Motor Function Classification System (GMFCS), Mini-Manual Ability Classification System (MiniMACS), Manual Ability Classification System (MACS), and Communication Function Classification System (CFCS) were used to classify participants.

**Results:** Assessments were well tolerated with 0-6.9% incomplete assessments. Floor or ceiling effects were not common with raw scores, age equivalents, or growth scales values but were frequently observed with scaled scores. Significant differences were found between the Bayley-4, GMFM-66, and PDMS-3 when participants were divided into groups by corresponding developmental domain classification scale. Strong associations (*p*<.05) were seen between all measures across Bayley-4 domains, PDMS-3 domains, and GMFM-66 total scores.

**Interpretation:** The Bayley-4, GMFM-66, and PDMS-3 are suitable for use in clinical practice and future clinical trials in *STXBP1*-RD.

## Introduction

*STXBP1-*related disorder (*STXBP1*-RD) is one of the most identified genetic causes of neurodevelopmental disorders and epilepsy. (1) Research to delineate the natural history of the condition has been largely performed through retrospective cohort studies. (2, 3) The largest study published included 534 individuals with *STXBP1*-RD and found severe global developmental delay, absent speech, autism, seizures, movement disorders, and a distinct tremor as common features. (4) Targeted therapies are emerging for *STXBP1-*RD (5), therefore a detailed prospective phenotypic description is needed.

A multisite prospective natural history study was recently initiated to evaluate individuals with *STXBP1*-RD longitudinally to choose appropriate clinical trial outcomes and better define the breadth of the condition. (6) Protocol development was completed jointly with patient advocacy organizations to determine the areas that are most important to measure. Qualitative interviews of caregivers through a disease concept model identified that gross motor function, communication, and decreased autonomy were common concerns. (7) Additionally, families also reported that communication difficulties, seizures, motor delays, and cognitive impairments are important concerns. (8)

There is limited information describing the gross motor, fine motor, and language function of individuals with *STXBP1*-RD using standardized measures. Because of the rarity of *STXBP1*-RD, using existing clinical assessments may be necessary due to the extensive resources needed to develop new condition-specific outcome measures. (9) Clinical outcome measures (COM) designed for children with cerebral palsy (CP) may be useful with individuals with *STXBP1*-RD because both conditions demonstrate many common features, including difficulty with gross and fine motor skills, communication difficulties, and abnormal tone. (10) Our previous work suggests that classification scales for children with CP can be reliably assessed in individuals with *STXBP1*-RD, (11) however, the validity of other commonly used COM has not been reported.

Another consideration when using COM is determining the best method of scoring. COM scoring may include raw scores, age equivalents (AE), growth scale values (GSV), and standard scores. Certain types of scores may be prone to demonstrate floor effects due to the level of functional impairment. Farmer and colleagues (12) reported that floor and ceiling effects were found when using age equivalents and norm referenced scores in individuals with rare genetic conditions. Difficulty measuring small changes in performance in COM towards the floor of a distribution and when using standard scores has been discussed in the literature concerning clinical trial readiness with developmental and epileptic encephalopathies (DEE). (13) Finally, Eisengart has advocated for the use of GSV, which are interval level data in rare pediatric conditions due to the potential for greater precision compared to ordinal scores (i.e., AE, raw scores). (14) Prior to the initiation of clinical trials for *STXBP1*-RD, there is a need to investigate the frequency of floor and ceiling effects with COM so that potential trial endpoints can be selected.

Many COM are designed to assess children within a specific age range while individuals with *STXBP1*-RD often have significant developmental delays so that measures designed for younger children may be developmentally appropriate. However, using COM intended for young children in older participants, risks ceiling effects in older children or adults with *STXBP1*-RD. Families have previously reported dissatisfaction with out of age range testing, as they felt that the COM did not accurately measure their child’s abilities. (15) Determining whether COM can detect small but meaningful changes in individuals with DEEs remains an area of active collaboration between researchers and patient advocacy groups. (16) Studies evaluating individuals with *STXBP1*-RD across age ranges are needed to evaluate the validity of COM performed within and outside their intended testing windows. Finally, potential interventional studies for *STXBP1*-RD should attempt to minimize the time required for testing to avoid participant fatigue affecting the assessments. By determining the relations between COM, informed decisions may be made regarding their inclusion or exclusion in clinical trials.

The aims of this study were to (1) describe the functional abilities of individuals with *STXBP1*-RD using COM assessing cognitive, expressive communication, receptive communication, fine motor, and gross motor function in a prospective natural history study, 2) quantify floor and ceiling effects as measured by raw scores, AE, GSV, and standardized scores, 3) explore the relations between and within COM.

## Methods

Participants were recruited to participate in a multisite prospective natural history study of *STXBP1*-RD (STARR: *STXBP1* Clinical Trial Ready; Clinical Trials Identifier: NCT06555965) (6), which was approved by the institutional review boards of all participating sites. Parents or legal guardians of all participants provided written informed consent. The selection of COM and classification scales for inclusion in this natural history study was made in consultation with the *STXBP1*-Disorders Foundation. (17)

Participants met the following inclusion criterion: a diagnosis of a *STXBP1*-RD, as determined through genetic testing as causative based on clinical and variant classification criteria. Exclusion criteria were: (1) a confirmed variant in a gene other than *STXBP1* known to contribute to a neurodevelopmental disability; (2) a significant disorder related to central nervous impairment; and (3) a history of birth at a gestational age less than 34 weeks, interventricular hemorrhage, structural brain deficit, or congenital heart disease. Data were analyzed from participants who were assessed from July 1, 2023 to June 30, 2026 who had completed at least one subtest of the Bayley Scales of Infant and Toddler Development, Fourth Edition (Bayley-4) Peabody Developmental Motor Scales, Third Edition (PDMS-3), and the Gross Motor Function Measure-66 (GMFM-66).

### Functional Classification Scales

The Gross Motor Function Classification System (GMFCS)(18), Mini-Manual Ability Classification System (Mini-MACS) (19), Manual Ability Classification System (MACS) (20) and Communication Function Classification System (CFCS) (21) were used to describe severity of functional impairment across their respective developmental domains. The GMFCS, Mini-MACS/MACS, and CFCS are 5-point ordinal scales designed to measure the gross motor, fine motor, and communication function, respectively, in children with CP, with lower levels indicating better ability. The GMFCS was completed in all individuals. The Mini-MACS was completed for individuals between 1 and 4 years while the MACS was used for individuals older than 4 years. The CFCS was completed for all participants at least 2 years old. While the MACS and GMFCS were developed for children under 18 years of age, participants over 18 years of age were also assessed using these measures. The Childhood Autism Rating Scale - Second Edition (CARS-2), which is a 15-item semi-structured assessment, was used to classify for risk for autism spectrum disorder (ASD) as no/low risk, mild to moderate risk, or severe risk. (22)

### Clinical Outcome Measures

The Bayley-4 is a norm-referenced test designed for children ages 16 days through 42 months. (23) The Bayley-4 consists of subtests assessing cognitive, expressive and receptive communication, fine motor, and gross motor function. Raw scores, GSV, and AE were calculated for all participants while scaled scores for cognitive, language, and motor function were calculated for children under 42 months of age. The PDMS-3 is a norm referenced test of motor development designed for children from birth through 72 months of age (24). The Hand Manipulation and Eye Hand Coordination subtests of the PDMS-3 were used. Raw scores and AE were calculated for all participants, while scaled scores were calculated for children under 72 months of age. The Gross Motor Function Measure-88 (GMFM-88) is an assessment of gross motor function for children with CP, from 5 months to 16 years of age. (25) A GMFM-66 score was calculated from items of the GMFM-88 using the Gross Motor Ability Estimator-3 software. (26) Floor and ceiling effects were defined as scoring the lowest or highest potential score respectively.

### Statistical analysis

Statistical analysis was completed using the R Studio. Chi-square goodness-of-fit tests were used to determine whether the distribution of classification scale levels differed from chance. Normality of data was evaluated using the Shapiro-Wilk test. All COM were non-normally distributed except for the Bayley-4 cognitive raw score and the GMFM-66. For the comparison of the normally distributed GMFM-66 scores across GMFCS levels, an ANOVA with Tukey post hoc comparisons was used. For non-normally distributed or ordinal data, Kruskal–Wallis tests were used with post-hoc pair-wise testing via Dunn’s test with Bonferroni adjustment for multiple comparisons. Kruskal-Wallis tests were used to determine whether raw scores, AEs, and scaled scores differed across the Bayley-4 domains when participants were classified by the corresponding scales (gross motor by GMFCS, fine motor by MiniMACS-MACS, and language by CFCS). Additionally, Kruskal-Wallis tests were used to evaluate differences in AEs across Bayley-4 domains. Pairwise comparisons between PDMS-3 Hand Manipulation and Eye Hand Coordination subtests were assessed using the Wilcoxon rank-sum test. Bivariate continuous relations were evaluated using Spearman rank correlation coefficients and were not corrected for multiple comparisons.

## Results

155 individuals with *STXBP1*-RD participated and consisted of 75 females and 80 males with a mean age of 8.2 (SD = 6.6 years) with a range of 0.4 to 31.2 years. 105 participants were evaluated using the CARS-2. Forty-two (40.0%) were classified as at either mild to moderate or severe risk for ASD while 21 (20.0%) were at classified as no/low risk for ASD. The classification scale levels among domains are presented in Table 1. Chi square Goodness of Fit analyses found statistically significant differences (*p* <0.001) from the expected distribution for the GMFCS (X^2^ = 41.0), MiniMACS-MACS (X^2^ = 41.7), and CFCS).

**Table 1:** Number of individuals with a classification score with percentage in parentheses.

|  | GMFCS<br>(N =155) | Mini-MACS/MACS<br>(N= 150) | CFCS<br>(N =134) |
| --- | --- | --- | --- |
| Level I | 29 (18.7%) | 7 (4.7%) | 4 (3.1%) |
| Level II | 61 (39.4%) | 53 (35.3%) | 12 (9.0%) |
| Level III | 18 (11.6%) | 38 (25.3%) | 30 (22.4%) |
| Level IV | 30 (19.4%) | 33 (22.0%) | 61 (45.5%) |
| Level V | 17 (11.0%) | 19 (12.7%) | 27 (20.1%) |

Abbreviations: CFCS = Communication Function Classification System; GMFCS = Gross Motor Function Classification System; MACS = Manual Ability Classification System; MiniMACS = Mini-Manual Ability Classification System

Bayley-4, GMFM-66, and PDMS-3 scores are presented in Table 2. Incomplete COM occurred in 0% of GMFM-66 and 1.3% of PDMS-3 assessments. Incomplete assessments with the Bayley-4 subtests occurred in 0-8.4%. The most common reason for incomplete testing was that the participant’s primary language spoken at home was not English and the assessor did not speak the caregiver’s primary language. Other reasons for incomplete testing included fatigue or behavioral/attention impairments. The frequency of floor and ceiling effects for the Bayley-4 domains, GMFM-66, and PDMS-3 domains varied according to scoring method. Floor effects were found in 0% of Bayley-4 domain raw scores and GSVs while ceiling effects occurred in less than 1.4% of assessments. Age equivalent scores demonstrated floor or ceiling effects in 0-2.8% of assessments across all domains. Floor effects were frequently observed in scaled scores (50.0-90.2% of assessments), but ceiling effects were not observed for any domain. For the GMFM-66, only one individual (0.6% of assessments) tested at the floor or ceiling score. Floor PDMS-3 raw scores in the Hand Manipulation and Eye Hand Coordination domains were found in 0.6% and 2.0% of assessments respectively while ceiling scores were not achieved in any participants. PDMS-3 AE scores were found at the floor value in 5.2% of Hand Manipulation and 9.8% of Eye Hand Coordination tests but were never found at the ceiling. PDMS-3 scaled scores were often at the floor value of each domain (74.7-86.5% of assessments) but were never found at the ceiling.

**Table 2:** Clinical Outcome Measures.

| Clinical Outcome Measure | Number | Number Unable to Complete (%) | Median | Minimum | Maximum | Number at Floor (%) | Number at Ceiling (%) |
| --- | --- | --- | --- | --- | --- | --- | --- |
| B4 Cognitive Total Score | 146 | 9 (6.2%) | 52 | 2 | 158 | 0 (0%) | 0 (0%) |
| B4 Cognitive Age Equivalent |  |  | 9 months | <20 days | > 41.08 months | 4 (2.7%) | 4 (2.7%) |
| B4 Cognitive GSV |  |  | 492 | 445 | 548 | 0 (0%) | 0 (0%) |
| B4 Cognitive Scaled Score | 38 | Not applicable | 1 | 1 | 4 | 32 (82.1%) | 0 (0%) |
| B4 Receptive Communication Total Score | 145 | 10 (6.9%) | 27 | 2 | 84 | 0 (0%) | 1 (0.7%) |
| B4 Receptive Communication Age Equivalent |  |  | 8 months | <20 days | > 41.08 months | 2 (1.4%) | 3 (2.1%) |
| B4 Receptive Communication GSV |  |  | 490 | 448 | 563 | 0 (0%) | 1 (0.7%) |
| B4 Receptive Communication Scaled Score | 37 | Not applicable | 1 | 1 | 12 | 23 (60.5%) | 0 (0%) |
| B4 Expressive Communication Total Score | 143 | 12 (8.4%) | 17 | 2 | 74 | 0 (0%) | 2 (1.4%) |
| B4 Expressive Communication Age Equivalent |  |  | 9 months | <20 days | > 41.08 months | 1 (0.7%) | 4 (2.8%) |
| B4 Expressive Communication GSV |  |  | 489 | 451 | 550 | 0 (0%) | 2 (1.4%) |
| B4 Expressive Communication Scaled Score | 37 | Not applicable | 1 | 1 | 9 | 19 (50.0%) | 0 (0%) |
| B4 Fine Motor Total Score | 145 | 10 (6.9%) | 37 | 3 | 91 | 0 (0%) | 0 (0%) |
| B4 Fine Motor Age Equivalent |  |  | 9 months | <20 days | > 41.08 months | 2 (1.4%) | 4 (2.8%) |
| B4 Fine Motor GSV |  |  | 497 | 451 | 556 | 0 (0%) | 0 (0%) |
| B4 Fine Motor Scaled Score | 38 | Not applicable | 1 | 1 | 7 | 25 (64.1%) | 0 (0%) |
| B4 Gross Motor Total Score | 155 | 0 (0%) | 70 | 2 | 116 | 0 (0%) | 1 (0.6%) |
| B4 Gross Motor Age Equivalent |  |  | 12 months | 20 days | > 41.08 months | 4 (2.6%) | 2 (1.3%) |
| B4 Gross Motor GSV |  |  | 502 | 451 | 552 | 0 (0%) | 1 (0.6%) |
| B4 Gross Motor Scaled Score | 40 | Not applicable | 1 | 1 | 3 | 37 (90.2%) | 0 (0%) |
| GMFM-66 Score | 155 | 0 (0%) | 51.3 | 0 | 100 | 1 (0.6%) | 1 (0.6%) |
| PDMS-3 Hand Manipulation Total Score | 155 | 0 (0%) | 16 | 0 | 98 | 1 (0.6%) | 0 (0%) |
| PDMS-3 Hand Manipulation Age Equivalent |  |  | 7 months | <1 month | 54 months | 8 (5.2%) | 0 (0%) |
| PDMS-3 Hand Manipulation Scaled Score | 75 | Not applicable | 1 | 1 | 8 | 56 (74.7%) | 0 (0%) |
| PDMS-3 Eye Hand Coordination Total Score | 153 | 2 (1.3%) | 14 | 0 | 83 | 3 (2.0%) | 0 (0%) |
| PDMS-3 Eye Hand Coordination Age Equivalent |  |  | 7 months | <1 month | 44 months | 15 (9.8%) | 0 (0%) |
| PDMS-3 Eye Hand Coordination Scaled Score | 74 | Not applicable | 1 | 1 | 7 | 64 (86.5%) | 0 (0%) |

Abbreviations: B4 = Bayley Scale of Infant and Toddler Development-Fourth Edition; GMFM-66 = Gross Motor Function Measure-66; PDMS-3 = Peabody Developmental Motor Scales Third Edition; GSV = Growth Scale Value

Bayley-4 scores by age across domains and types of scoring are shown in Figure 1. Scatterplots show individual participant scores versus age for Bayley-4 domains across each score type. Total, AE, and GSV scores increased with chronological age across all domains. Scaled Scores show floor effects and declined with age reflecting an increased divergence from age-expected performance over time.

**Figure 1.**
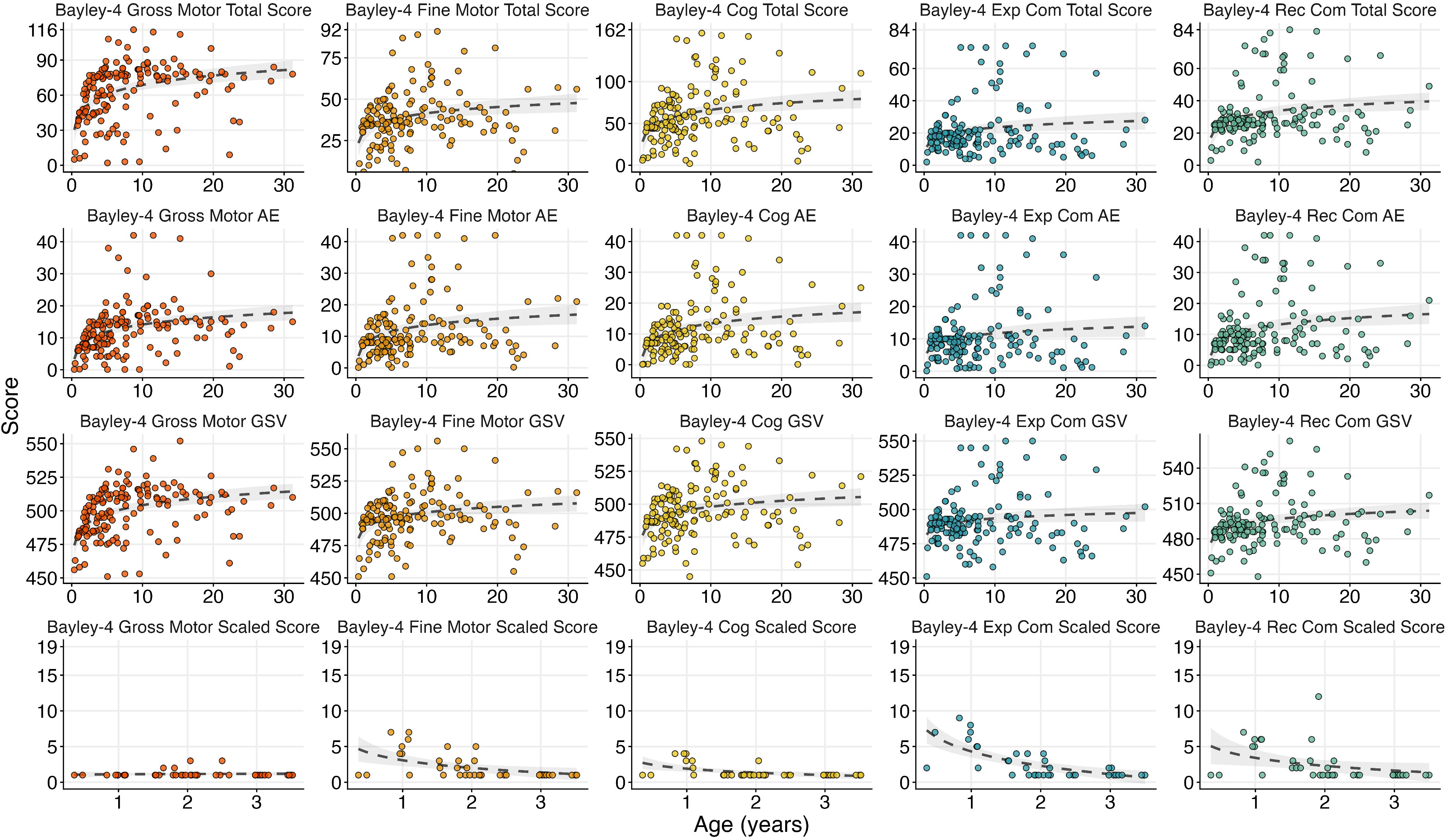
Bayley-4 scores by age across domains and types of scoring. Scatterplots show individual participant scores versus age (years) for Bayley-4 domains across each score type: Total (raw) Score, Age Equivalent (AE) score in months, Growth Scale Value (GSV), and Scaled Score. Dashed lines represent logarithmic fitted trend lines with shaded 95% confidence bands. Total, Age Equivalent, and Growth Scale Value scores generally increased with chronological age across all domains. Scaled Scores, which are calculated for children less than 42 months of age and are normed relative to typically developing peers, show floor effects and declined with age reflecting an increased divergence from age-expected performance over time. Abbreviations: AE = Age Equivalents; Bayley 4 = Bayley Scales of Infant and Toddler Development-Fourth Edition; Cog = Cognitive; Ex Com = Expressive Communication; GSV = Growth Scale Values; Rec Com = Receptive Communication

Median AE scores for Bayley-4 domains ranged from 8 months (receptive communication) to 12 months (gross motor). A significant difference in AE’s was found when comparing across Bayley-4 domains (*p* < 0.001). Post hoc testing found that gross motor scores were higher than expressive communication, receptive communication, and cognition while fine motor was significantly higher than expressive communication (*p* < 0.05) (Figure 2). All other comparisons did not reach statistical significance. The median AEs for the PDMS-3 Hand Manipulation and Eye Hand Coordination domains were 7 months. There was no significant difference between AE for the PDMS-3 Hand Manipulation and Eye Hand Coordination domains.

**Figure 2.**
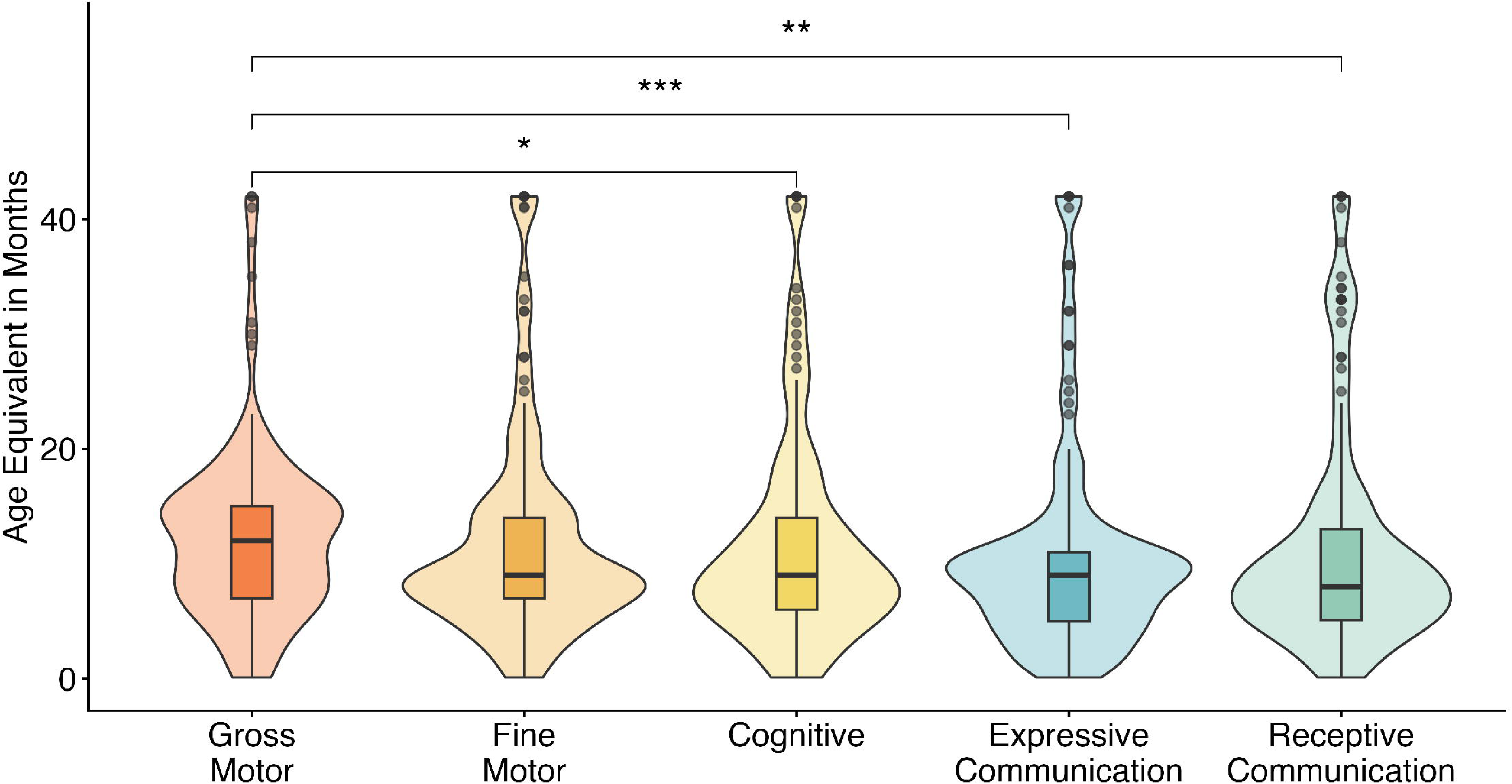
Comparison of age-equivalent scores across Bayley-4 domains. Violin plots with overlaid boxplots show age-equivalent scores (months) across Bayley-4 domains. Boxplots represent the median and interquartile range; violin outlines depict the full distribution of scores within each domain. Horizontal brackets indicate statistically significant pairwise comparisons between domains (* = *p* < 0.05, ** = *p* < 0.01, *** = *p* < 0.001). Gross Motor scores were significantly higher than Fine Motor, Expressive Communication, and Receptive Communication scores.

Significant differences were observed when comparing assessments of gross motor skills, Bayley-4 gross motor domain and GMFM-66 across GMFCS levels. Similarly, there were significant differences when comparing measures of fine motor skills, Bayley-4 and PDMS-3 Hand Manipulation and Eye-Hand Coordination across MiniMACS-MACS classifications. There were also significant differences observed when comparing Bayley-4 receptive communication and expressive communication domain scores across CFCS classifications. These differences were significant for all score types (*p* < 0.01) except scaled scores. Figure 3 shows post-hoc pair-wise comparisons between severity levels.

**Figure 3.**
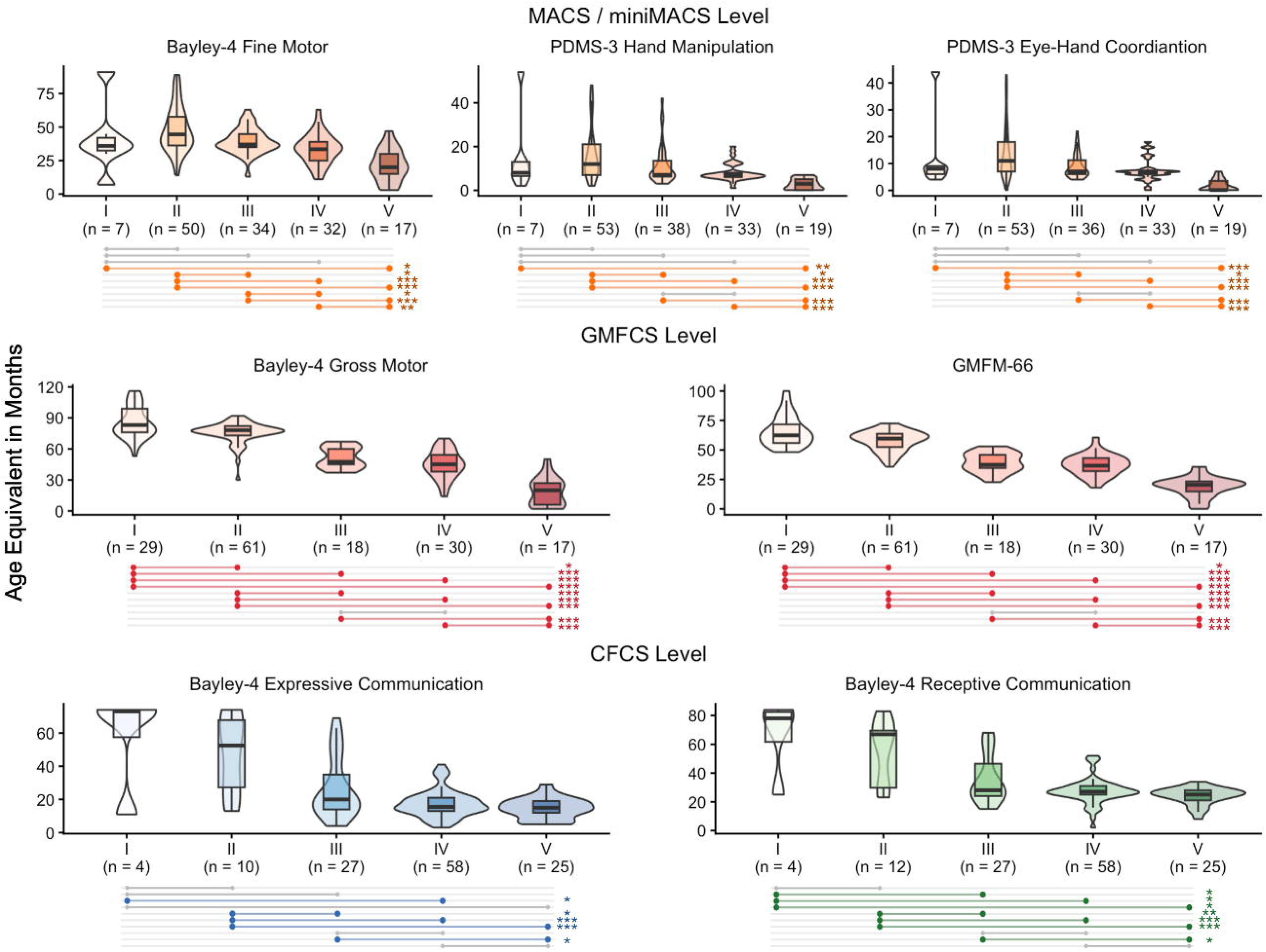
Association between functional classification levels and age-equivalent clinical outcome measure scores. Violin plots with overlaid boxplots show age-equivalent scores (months) for clinical outcome measures, stratified by classification level (I–V), with sample sizes (n) indicated below each group. Boxplots represent the median and interquartile range; violin outlines depict the full distribution of scores within each level. Horizontal lines below each plot indicate pairwise comparisons between levels, with colored dots marking statistical significance (* = *p* < 0.05, ** = *p* < 0.01, *** = *p* < 0.001) after correction for multiple comparisons. Across all panels, higher functional classification levels (indicating greater impairment) were associated with lower age-equivalent scores. Abbreviations: Bayley 4 = Bayley Scales of Infant and Toddler Development-Fourth Edition; CFCS = Communication Function Classification System; GMFCS = Gross Motor Function Classification System; GMFM-66 = Gross Motor Function Measure-66; MACS = Manual Ability Classification System; MiniMACS = Mini-Manual Ability Classification System; PDMS-3 = Peabody Developmental Motor Scales -Third Edition

There were strong correlations between domains from the same COM across score types, except scaled scores (rho values = 0.52-0.94, p>0.001; Figure 4). There were also strong correlations between different COM evaluating the same functional domain. The correlation between the GMFM-66 and Bayley-4 gross motor domain raw score was 0.94, while correlations between the raw scores of the Bayley-4 fine motor domain and the PDMS-3 hand manipulation and eye hand coordination subtests were 0.85 and 0.81, respectively.

**Figure 4.**
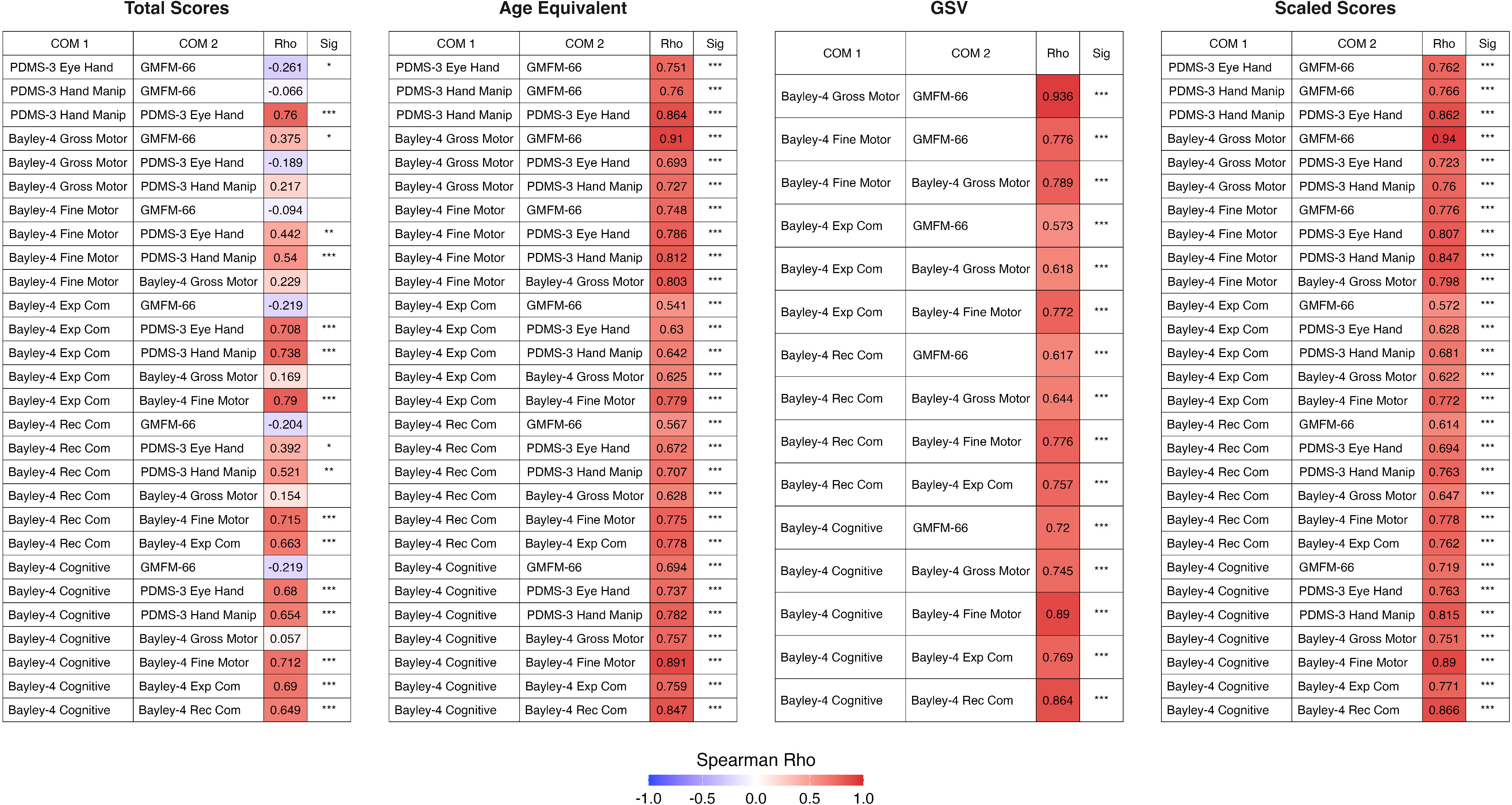
Spearman correlations between clinical outcome measures across score types. Tables show pairwise Spearman correlation coefficients (rho) between Bayley-4 subdomain scores and corresponding PDMS-3/GMFM-66 measures, calculated separately for four scoring metrics: Age Equivalent, Growth Scale Values, Scaled Scores, and Total Scores. Cell shading reflects the strength and direction of the correlation. Asterisks denote statistical significance (* = *p* < .05, ** = *p* < .01, *** = *p* < .001). Most measures were strongly and positively correlated across metrics, except for Scaled Scores, which show weak or negative, non-significant correlations with several measures. Abbreviations: Bayley 4 = Bayley Scales of Infant and Toddler Development-Fourth Edition; Ex Com = Expressive Communication; Eye Hand = Eye Hand Coordination; GMFM-66 = Gross Motor Function Measure-66; GSV = Growth Scale Values; Hand Manip = Hand Manipulation; PDMS-3 = Peabody Developmental Motor Scales-Third Edition; Rec Com = Receptive Communication.

## Discussion

Our study reports the most comprehensive description of gross motor, fine motor, cognitive, and language function in a prospective natural history study of individuals with *STXBP1*-RD using COM. Individuals with *STXBP1*-RD demonstrate marked developmental delays across all domains and most often present with skills equivalent to those between a 7-to 12-month-old level, regardless of chronological age. By using standardized COM, we were able to more carefully define the degree of developmental delay compared to previous reports of *STXBP1*-RD. (2–4) Our work provides insight into the feasibility of applying natural history protocols using standardized COM for genetic developmental and epileptic encephalopathies more broadly and provides a framework for protocol design for conditions of similar developmental severity.

Our results demonstrate that the Bayley-4, GMFM-66, and PDMS-3 are suitable for use in clinical practice and future clinical trials with *STXBP1*-RD. All assessments were well tolerated as evidenced by high rates of completion across multiple sites and infrequent floor or ceiling effects when measured using raw scores, AE, or GSV, suggesting that the full phenotype of motor, language, and cognitive function in *STXBP1*-RD can be assessed using these COM. These tests were completed regardless of the age of participants, thereby showing that out of age range testing may be tolerated by participants with *STXBP1*-RD despite the previously reported concerns. (15)

Our investigation did not attempt to standardize the order of testing administration since testing was often completed in the context of physical therapy, occupational therapy, and clinical psychology evaluations so that the order of COM testing could be adjusted to complete COM. Future clinical trials could attempt to further improve completion rates and avoid fatigue by carefully considering the order of assessments to prioritize the COMs which that are the primary outcomes for a specific disorder. One consideration for our high rates of COM completion and infrequent floor effects is that participants who are unlikely to be able to complete testing due to behavioral issues, or those who would be more likely to demonstrate a floor effects, might also be less likely to be able to travel to participate in the study. The development of remote assessment tools of gross motor, fine motor, and language function may be necessary to maximize inclusion of all individuals with *STXBP1*-RD in future studies and clinical trials.

The validity of Bayley-4, GMFM-66, and PDMS-3 was also supported by the significant differences found when participants were classified using the relevant classification scales. When examining AEs across Bayley-4 domains, language function was more affected than gross motor function, a finding supported by previous studies. (3, 4) By using previously developed COM rather than developing *STXBP1*-RD specific measures, researchers and clinicians will avoid the time and expense of developing and validating new measures. By combining GMFCS age bands and the MiniMACS and MACS together due to limited numbers of participants in some age groups, differences between classification levels could also be more difficult to detect. For example, a GMFM-66 score of 50 may occur in a child who is GMFCS Level I if they are 18 months old, while a child who is 6 years old would have higher GMFCS classification with that same score. Future clinical trials may also limit the age range of participants as part of their study’s inclusion criteria, therefore additional analysis of different age bands with larger numbers of participants may be necessary to facilitate clinical trial readiness.

Our findings suggest that scaled scores should be avoided due to frequent floor effects, weak correlations with other measures, and the inability to differentiate between classification scale levels. Our findings are like other studies that have reported that scaled scores are not able to detect change over time. (12) Future reports of longitudinal data and investigations of the responsiveness of the Bayley-4, PDMS-3, and GMFM-66, are necessary to determine the validity of these tools in future clinical trials for *STXBP1*-RD.

Correlational analysis found strong relations when comparing within functional domains of the Bayley-4, GMFM-66, and PDMS-3 when using raw scores, GSV, and AE, which suggests that these COM are measuring gross and fine motor function validly. Strong correlations between the Bayley-4 gross motor subscale and GMFM-66 and between the PDMS-3 subscales and Bayley-4 fine motor domain suggest that clinicians or researchers who are trying to minimize the time required for testing may decide to use only one of each correlated pairs of COM. However, the minimal important clinical differences for the Bayley-4, GMFM-66, and PDMS-3 for individuals with *STXBP1*-RD have not been determined, so it is unknown which assessment is the optimal tool. Future studies examining COM feasibility and validity with larger samples sizes within specific age ranges are necessary to facilitate clinical trial readiness since interventional trials may target specific age ranges.

## Conclusions

We provide evidence that individuals with *STXBP1*-RD demonstrated high completion rates for the Bayley-4, the GMFM-66, and the PDMS-3. Thus, these tests are well tolerated by this patient population. Indeed, the lack of floor or ceiling effects suggests that the phenotypic spectrum of motor, language, and cognitive function in *STXBP1*-RD can be assessed using these COM. The validity of these tests was further supported by high correlations when comparing within functional domains, and by the significant differences found when participants were classified using relevant scales. We conclude that the Bayley-4, GMFM-66, and PDMS-3 are suitable for use in clinical practice and future interventional trials for *STXBP1*-RD.

## Data Availability

All data produced in the present study are available upon reasonable request to the authors

## Acknowledgements

We would like to thank the following individuals for their contributions to this project:

<u>Stanford University</u>

Amy Weisman, Nicolle Kaytsner, Sophia Magana, Sarah Niswonger, Christine Hedugus, Stephanie Bragg, Sweta Patnaik, Rayann Solidum, Julie Nemerson, Prathy Teeyagura Arushi Gehani

<u>Baylor College of Medicine and Texas Children’s Hospital:</u>

Alvina Zia, Ekaterina Sanchez-Romero, Isaiah Valentine, Elaine Seto, Danielle S. Takacs, Sruthi Thomas, Roberta Olivares, Natasha Feuerbach, Kristen S. Fisher, Mikael Guzman-Karlsson, Arden Wheeler, Andres Jimenez-Gomez, Maheen Rizvi, Karina Henriquez, Alysson Nillas, and Hyunmin Shin.

<u>Weill Cornell Medicine</u>

Natalie Wayland, Amelia Stone, Natasha Basma, Aida Osis, Tyler Demes, Dara Jones, Ji-Sun Kim, Jen Cross

<u>Children’s Hospital Colorado</u>

Megan Stringfellow, Kaitlyn Kennedy, Kiara Hamlin, Margarita Saenz, Katie Angione, Dana Bennink, Andrea Gerk, Julie Vanek, Brittany Gladfelter, Ann Reynolds, Kristie Malik, Kourtney Santucci, Kelly Bradley, and Ryleigh VandenBroeke

